# POPULATION-BASED SERO-EPIDEMIOLOGICAL STUDY PROTOCOL FOR THE IMPACT OF SMOKING ON SARS-COV-2 INFECTION AND COVID-19 OUTCOMES – THE TROINA STUDY

**DOI:** 10.1101/2021.04.29.21256236

**Authors:** R. Polosa, V. Tomaselli, P. Ferrara, A. C. Romeo, S. Rust, D. Saitta, F. Caraci, C. Romano, M. Thangaraju, P. Zuccarello, J. Rose, M Ferrante, J. Belsey, F. Cibella, E. Interlandi, R. Ferri

## Abstract

After the global spread of severe acute respiratory syndrome coronavirus 2 (SARS-CoV-2), research has highlighted several aspects of the pandemic, focusing on clinical features and risk factors associated with infection and disease severity. However, emerging results on the role of smoking in SARS-CoV-2 infection susceptibility or COVID-19 outcomes are conflicting, and their robustness remains uncertain. In this context, this project aims at quantifying the proportion of SARS-CoV-S antibody seroprevalence, studying the changes in antibody levels over time, and analyzing the association between smoking status and infection using seroprevalence data.

The added value of this research is that the current smoking status of the population to be studied will be biochemically verified, in order to avoid the bias associated with self-reported smoking status. As such, the results from this survey may provide actionable metric to study the role of smoking in SARS-CoV-2 spread, and therefore implement the most appropriate public health measures to control the pandemic.

The research design involves a 6-month prospective cohort study with serial sampling of the same individuals. Each participant will be surveyed about their demographics and COVID-19-related information, and blood sampling will be collected upon recruitment and at specified follow-up time points (namely, after 8 and 24 weeks). Blood samples will be screened for the presence of SARS-CoV-2 specific antibodies and serum cotinine.

Overall, we expect to find a higher prevalence of antibodies in individuals at high-risk for viral exposure (i.e., healthcare or other essential workers), according to previous literature, and to refine current estimates on the association between smoking status and SARS-CoV-2/COVID-19. Our results may serve as a reference for future clinical research and the methodology could be exploited in public health sectors and policies.

## Scientific Background

Severe acute respiratory syndrome coronavirus 2 (SARS-CoV-2) is the novel coronavirus strain that was first reported as a cluster of viral pneumonia cases of unknown etiology in Wuhan, the capital city of the Hubei Province in China on December 31, 2019. The spreading of SARS-CoV-2 reached pandemic proportion in March 2020, and as of the April 2021 more than 141 million cases had been confirmed, and more than 3 million fatalities had occurred worldwide (1).

In late February 2020, the first non-imported cases of COVID-19 were identified in Italy. Since then, SARS-CoV-2 spread rapidly to the community, as reported by national health authorities (2). On May 23, 2020, at the time of project proposal design, the Italian Ministry of Health reported that of the 229,327 people who had contracted the virus, 57,752 were still positive (of whom 8,695 were hospitalised with symptoms - 15%; 572 were admitted in intensive care units - 1,0%; and the remaining 48,485 were self-isolating at home - 84%), 32,735 had died (14.3%) and 138,840 healed (60.5%) on a total of 2,164,426 tested cases (3). As of April 2021, 3,870,131 cases and 116,927 deaths have been recorded in the country.

Since the beginning of the pandemic, the global scientific community was engaged in intense research efforts to understand all aspects of this public health crisis, from the clinical features of the disease and risk factors for adverse outcome, the patterns of viral spread in the population, the role of asymptomatic or subclinical cases in human-to-human transmission, and the serological response. The clinical features of the Novel Coronavirus Disease 2019 (COVID-19) range from a self-limited flu-like syndrome to progressive lung involvement with respiratory failure and widespread systemic effects (4-6). Epidemiological surveillance has primarily focused on hospitalized patients with severe disease, and, as such, the full spectrum of the disease, including the extent and proportion of mild or asymptomatic infections is less clear. Evidence suggests that asymptomatic or oligo-symptomatic infection is not uncommon (7-10). A recent review reported that at least one third of SARS-CoV-2 infections are asymptomatic and that almost three quarters of persons who are asymptomatic at the time of a positive PCR test result will remain asymptomatic (11). Remarkably, sequalae of previous viral pneumonia have been reported in chest CT scans of asymptomatic individuals (12) and SARS-CoV-2 transmission from asymptomatic cases to others has been documented (12,13).

In this frame, a seroprevalence study is an ideal attempt for measuring the true infection rates in general or specific populations (10,14,15) In Italy, a nationwide survey was conducted by the National Institute of Statistics (ISTAT), reporting an IgG seroprevalence of 2.5%. (17). However, there is limited use of seroprevalence studies in order to retrospectively identify potential predictors of infection susceptibility and disease severity, both in the general population and in specific sub-groups (such as smokers, pediatric populations, elders).

Several risk factors for severe COVID-19 have been identified, including cardiovascular disease, diabetes, obesity, and COPD (18-21). Intuitively, one important additional risk factor is expected to be cigarette smoking. Smokers have a higher risk for developing viral and bacterial respiratory infections (22-24), being five times more likely to have influenza and twice more likely to develop pneumonia (25).

However, the role of smoking in SARS-CoV-2 infection susceptibility or COVID-19 outcomes is still unclear. While there appears to be a higher risk for IU admission and adverse outcomes (26-28), it has been reported that the prevalence of smoking among hospitalized COVID-19 patients is far lower than would be expected based on population smoking prevalence (29-32). These findings were initially derived from Chinese case series, and while it is possible that the prevalence of smokers in Chinese case series may be underrepresented due to inaccurate recording of their smoking status, similar findings have been reported in France (30), Germany (33), Italy (34), and in the US (31,32).

It is not clear how far the underrepresentation of smokers among COVID-19 inpatients reflects problems with poor reporting of the smoking status. Given the challenging circumstances of the pandemic, recall or reporting bias cannot be excluded. The possibility for inaccurate recording, false-reporting or underreporting of the smoking status due to the extremely challenging situations at wards/ICUs with work overloads and operating in a persistent state of emergency should not be underestimated. Improving the quality of clinical and behavioral data mandates the need for an accurate and dedicated recording of the smoking status. Alternatively, population-level data collected outside of hospital settings are required. Another important limitation is that most of the observations were unadjusted for smoking-related comorbidities, which are known to be associated with higher risk for an adverse outcome in COVID-19 patients (18). Lack of adjustment for relevant confounders means it is not possible to disentangle the effect of smoking. Addressing all these limitations is important for evaluating clinical risk, developing clear public health messages, and identifying targets for intervention.

### Research Objectives

Surveillance of antibody seropositivity in a population can allow inferences to be made about the cumulative incidence of infection in the population. Additionally, little is currently known about antibody kinetics. Asymptomatic infected persons may clear the virus more quickly than do symptomatic patients, and antibody titers in the former are likely to be lower, if they seroconvert at all, than in infected symptomatic patients (35). Furthermore, understanding the association between smoking and SARS-CoV-2 infection susceptibility or COVID-19 outcomes is generally limited and of poor quality.

To summarize, there is a need for robust, population-based evidence on the association of smoking with SARS-CoV-2 infection and COVID-19 outcomes, adjusting for potential confounding variables (e.g., sociodemographic characteristics, key worker status, and comorbid health conditions), and a population seroprevalence study could be useful for this goal. The following key research questions will be addressed:

1. Does smoking increase susceptibility to SARS-CoV-2 infection?
2. Does smoking increase susceptibility to COVID-19 outcomes?
3. Does smoking affect the serological response after SARS-CoV-2 infection?

### Research Proposal

We propose a 6-months prospective study using in combination a random population sample (taken from residents of the town of Troina; the town with the highest prevalence of positive SARS-CoV-2 cases in Sicily) and a convenience sample (taken from staff of the Troina’s main health care establishment), to investigate the prevalence of past infection, as determined by seropositivity (anti SARS-CoV-2-specific IgG by ELISA). The biochemically-verified smoking status of the study population will be correlated with serological data and COVID-19 outcomes (i.e., clinical symptoms and hospitalization).

### Specific Aims

Epidemiological exposure data and venous blood (for measurements of anti-SARS-CoV-2-specific IgG and serum cotinine levels) will be systematically collected. Demographic, medical history and epidemiological exposure data will be recorded from specifically designed questionnaires (for COVID-19 outcomes, relevant comorbidities, and smoking status) and shared rapidly in a format that can be easily aggregated, tabulated and analyzed across many different regional (or national and international) settings for timely estimates of COVID-19 virus infection and its immunologic response rates according to the smoking status, as well as to inform public health responses and policy decisions.

In summary, the main objectives of the study will be to:

1. Quantify the proportion of SARS-CoV-2 infected people (by serology assessment of anti-SARS-CoV-2 IgG levels) in a random population sample at baseline;
2. Quantify the proportion of SARS-CoV-2 infected people (by serology assessment of anti-SARS-CoV-2 IgG levels) in a convenience sample (hospital staff) at baseline;
3. Quantify the proportion of participants with asymptomatic SARS-CoV-2 infection;
4. Quantify the proportion of mildly-to-moderately symptomatic (management at home) COVID-19 cases among all participants;
5. Quantify the proportion of severely symptomatic (management at hospital) COVID-19 cases among all participants;
6. Quantify the proportion of biochemically verified (by serum cotinine) current, former and never smokers among all participants (random population sample + convenience sample);
7. Quantify the proportion and analyze the association between smoking status (current, former and never smokers) and:
  a. SARS-CoV-2 infection (asymptomatic and symptomatic combined)
  b. asymptomatic SARS-CoV-2 infection
  c. mild-to-moderate COVID-19 disease (management at home)
  d. severe COVID-19 disease (hospitalization needed)
8. Record relevant clinical confounders known to be associated with COVID-19 outcomes;
9. Compare anti-SARS-CoV-2 IgG levels at baseline among current, former and never smokers;
10. Monitor and compare anti-SARS-CoV-2 IgG levels changes from baseline to 2-months and 6-months among current, former and never smokers.

## Methods

### Study design

This study will investigate the association between biochemically verified smoking status of the study population and serological data as well as COVID-19 outcomes (i.e., clinical symptoms and hospitalization). The research design involves a 6-month prospective multiple cohort study with serial sampling of the same individuals each time. Sampling will be commenced in July and repeated at 8 weeks (Figure 1). A final follow-up visit will be carried out at 24 weeks.

### Study population

Within the geographic scope of the study, high incidence of positive cases has been identified in the general population of the town of Troina, having the highest prevalence of positive SARS-CoV-2 cases in Sicily according to the surveillance data on COVID-19 cases available from the Assessorato alla Salute della Regione Sicilia. The study population will consist of a population-based, age-stratified cohort in Troina that will be sampled through random selection of town residents. Identification and recruitment of participants will expand over different age groups in order to determine and compare age-specific attack rates. For logistic reasons, specimen and data collection will be performed at a single location, asking participants to travel to that location to participate in the study. Targeted testing will also be extended to a convenience sample consisting of about 600 staff members of Troina’s main health care establishment (high risk individuals).

### Eligibility criteria

Any individual identified for recruitment, irrespective of age, can participate. Exclusion criteria will be refusal to provide informed consent, or contraindication to venipuncture. Suspected or confirmed active/acute or prior SARS-CoV-2 infection should not be considered as an exclusion criterion for this investigation. Doing so would underestimate the extent of infection in the population. For individuals currently receiving medical care for COVID-19 infection, a family member or proxy may be used to complete the questionnaire on his/her behalf.

### Smoking status definition

Current smokers will be defined as those who report that they smoke and have serum cotinine levels ≥ 20 ng/mL. Former smokers will be defined as those who report that they used to smoke in the past but not now and have serum cotinine levels < 20 ng/mL. Never smokers will be defined as those who report that they never smoked smoke in the past and have serum cotinine levels < 20 ng/mL.

### Data collection

Each participant recruited into the study will be asked to complete a questionnaire which will record the following information: demographics, information about known COVID-19 disease and relevant clinical course, comorbidities, and smoking status (see attached).

### Specimen collection

A small amount of blood (10 mL) will be collected from each participant upon recruitment (T0) and at specified follow-up time points (T8wks, T24wks).

Specimen transport and Biobanking: For each biological sample collected, the time of collection, the conditions for transportation and the time of arrival at the study laboratory will be recorded. Specimens should reach the laboratory as soon as possible after collection. Serum should be separated from whole blood and stored at -20°C or lower and shipped on dry ice. A biobanking facility will be established in Troina.

### Sample storage

Prior to testing, serum samples will be stored at -80°C at the reference biobanking facility. It is recommended to aliquot samples prior to freezing, to minimize freeze thaw cycles.

### Serological testing

Serologic assays of high sensitivity and specificity for SARS-CoV-2 have been recently validated and published. Serum samples will be screened for the presence of SARS-CoV-2 specific antibodies using a quantitative enzyme linked immunosorbent assay (ELISA) test for anti-SARS-CoV-2 IgG (Euroimmun, CND W0105040619). Serum samples will be stored at -80°C until use. and the assay will be performed according to manufacturer’s protocol. The neutralization capability, specificity and sensitivity of the test have been thoroughly investigated and recently published together with assay validation (Okba et al. 2020). Reagent wells of the assays are coated with recombinant structural protein (S1 domain) of SARS-CoV-2. The optical density (OD) will be detected at 450 nm, and a ratio of the reading of each sample to the reading of the calibrator, included in the kit, will be calculated for each sample (OD ratio). The cut-off value for IgG OD ratio is 0.3. Following blocking, diluted serum (1:100 or 2-fold serially diluted for titers) will be added and incubated at 37°C for 1h in the 96-well microtiter ELISA plates. Antigen-specific antibodies will be detected using peroxidase-labeled rabbit anti– human IgG (Dako, https://www.agilent.com) and TMB as a substrate. The absorbance of each sample will be measured at 450 nm. Laboratory procedures involving sample manipulation must be carried out in a biosafety cabinet.

### Cotinine Assay

About 1mL of serum will be pipetted into 10 ml tubes and 100 ng/mL of Ortho-cotinine used as internal standard will be added. Then, about 50µL of 0.1M aqueous sodium hydroxide solution will be added to the culture tube followed by 325µL of Chloroform. The tube will be secured with cap and Vortex mixed for ∼3 minutes (using VX 2500 Multi Tube Vortex Mixer) and centrifuged for ∼4 minutes (in Beckmann Allegra centrifuge) at 2500 rpm. Using a glass Pasteur Pipette, the top aqueous layer will be removed and discarded into the hazardous waste container and the organic layer will remain in the tube. About ∼100mg (0.1g) of anhydrous sodium sulfate will be added to the organic layer and allowed to rest for ∼3 minutes (which will allow the sodium sulfate to absorb any water that may be present in the organic layer). In the end, the clear organic layer (with no water) will be carefully removed (without disturbing the settled sodium sulfate), concentrated to ∼100 µL vial insert and placed in a GC vial. The concentrated sample will be capped and arranged on to auto sampler tray for GC injection. One microliter (µL) of each sample will be injected into HP-5 Capillary GC Column (0.32 mm ID, 25 m length and 0.52µm film thickness; bonded 5% phenyl and 95% dimethylpolysiloxane) of GC-NPD. The inlet temperature will 250°C at split-less mode. The initial oven temperature will be 70°C with 1-minute hold and then increased to 230°C at the rate of 25°C/minute. Every batch of sample will be run with 6 calibration levels (20, 50, 100, 200, 400, 600 ng/mL), 4 quality controls (20, 100, 400, 600 ng/mL) and 1 blank control for accurate quantification. The amount of cotinine will be reported in ng/mL The limit of quantification (LOD) of cotinine is 20 ng/mL.

### Statistical Considerations and Statistical Plan

#### Sampling design

Estimates of margin of error as a function of seroprevalence are low for 300 samples. We will be aiming for > 1000 samples of a representative sample of the population by gender and age groups (0-17 years, 18-65 years, and 66 years and over).

For the enrolment of participants into the study the following inclusion and exclusion criteria need to be fulfilled.

#### Inclusion in population criteria

⍰ Aged into the above groups
⍰ Living in the village of interest
⍰ Willing to participate in the baseline and to be re-contacted for follow-up waves

#### Exclusion from population criteria

⍰ Persons belonging to the institutionalized population

#### Exclusion from sample (censoring) criteria

⍰ Moved to another village during the study period
⍰ Those stating explicitly their wish not to be re-contacted for this study

In order to correctly represent the population involved, the age groups of interest have been assigned to three different categories by gender in terms of population size. A corresponding targeted sample size for this study is specified for each category.

In the entire population, we estimated several sample sizes depending on the margin of error equal to 0.03, 0.04, or 0.05 for estimate proportions of sampled population, as shown in the following table:

| TROINA |  |  |  |  |  |
| --- | --- | --- | --- | --- | --- |
|  |  |  | *Margin of error*<br>*3%* | *Margin of error*<br>*4%* | *Margin of error*<br>*5%* |
| Age groups | Gender | N | (N*1308) /<br>TOTAL 9094 = n | (N*613) /<br>TOTAL 9094 = n | (N*384) /<br>TOTAL 9094 = n |
| 0-17 y. | Male | 694 | 100 | 47 | 29 |
|  | Female | 648 | 93 | 44 | 27 |
|  | **Total** | **1342** | **193** | **90** | **57** |
| 18-65 y. | Male | 2751 | 396 | 185 | 116 |
|  | Female | 2803 | 403 | 189 | 118 |
|  | **Total** | **5554** | **799** | **374** | **235** |
| 66 + years | Male | 961 | 138 | 65 | 41 |
|  | Female | 1237 | 178 | 83 | 52 |
|  | **Total** | **2198** | **316** | **148** | **93** |
| TOTAL | Male | 4406 | 634 | 297 | 186 |
|  | Female | 4688 | 674 | 316 | 198 |
|  | **Total** | **9094** | **1.308** | **613** | **384** |

We will draw a multi-layered sample with a confidence level of 0.97% in order to minimize the margin of error to 3% and ensure the best reliability of the sample data. The planned total sample size comprises up to 1,308 participants at recruitment stage. The attrition rate is estimated at 10%.

### Reporting of findings

Information will be collected in a standardized format according to the questionnaires and tools in the protocol (see attached). The data shared should include only the study identification number and not any personal identifiable information. We will report the following information:

1. the number of households and the number of individuals included;
2. the age and sex of all individuals included
3. the antibody levels in the sample of all individuals included
4. the number of individuals with serologic evidence of COVID-19 virus infection (stratified by age.)
5. the number of individuals with serologic evidence of COVID-19 virus infection who have reportedm symptoms.
6. the number of individuals with known previous SARS-CoV-2 infection who were hospitalized or recovered at home.
7. the number of individuals with serologic evidence of COVID-19 virus infection who are current, former and never smokers (biochemically verified by serum cotinine assay).

### Statistical analysis plan

Socio-demographic and baseline characteristics will be summarised for the Troina population, the convenience sample and for the total sample recruited. Categorical variables will be reported as numbers and proportions +/-95% CI. Between-groups comparisons will be carried out using chi-square testing or Fisher’s exact test, as appropriate. Continuous variables will be reported as means and standard deviations and as medians and interquartile ranges. Between-groups comparisons will be carried out using tests like ANOVA, Mann-Whitney U test or Wilcoxon-signed rank test, as appropriate.

The proportions of patients developing a positive PCR test for SARS-CoV-2 result over the course of the study will be reported as numbers and proportions +/-95% CI, separated by subgroups identifying those with symptomatic or asymptomatic infection.

Data on change in antibody levels from baseline to follow-up will be presented for the whole recruited population. Statistical significance of change will be estimated using repeated-measures t-testing

Correlation between smoking status and the risk of COVID-19 infection will be tested using a 3×2 Chi-square Test of Independence.

A p-value <0.05 will be considered to represent the threshold of statistical significance for all comparisons

### Data collection procedures

In order to accomplish the specific aims of the project, data will be stored in a database system, after each completed collection step, and transfer to central data management for further data processing and merging.

Each participant will be assigned a unique identification code consisting of a number identifier which will be used for data merging. After data collection, data validation checks will be performed as agreed in a data validation plan, and data cleaning procedures will be used as applicable to ensure best achievable quality of data for analysis purposes. Only the study staff working with the fieldwork provider will be able to identify the participants based on the identification codes.

Only de-identified data will be transferred from the field work provider to the central data management. All electronic data files will be kept on secure servers with backup processes in place. Personal data of participants must be strictly kept separately from study data and is accessed only by authorized staff for the purposes of the study conduct.

Before starting data collection, the involved staff will receive training on the background and objectives of this study, on eligibility criteria, on the participant selection procedure, on ethical obligations, on completion and validation of the procedure, as well as on the data collection platform. Local field work staff will be trained for each relevant data collection process and logistic related procedure.

Eligible subjects are informed about the study purpose, their requested tasks, time of involvement, data confidentiality and data protection. Once they stated their willingness to participate, they will proceed with the screening and study enrolment. Enrolled subjects have the right to stop their participation at any time without any penalty. Preferably, the reason for premature end of participation will be recorded. Participants who drop-out will not be replaced. Every effort will be made to protect participant confidentiality according to the GDPR.

### ETHICAL AND CONFIDENTIALITY CONSIDERATIONS

#### Research Ethics Committee (REC) review

- Before the start of the study, approval will be sought from the local REC (Comitato Etico IRCCS Sicilia - Oasi Maria SS) for the study protocol, informed consent forms and other relevant documents e.g. advertisements and GP information letters
- Substantial amendments that require review by REC will not be implemented until the REC grants a favorable opinion for the study
- A final report with the results will be submitted to the REC within 90 days of the date on which the study is declared ended

### Participants’ Consent

The Principal Investigator (PI) will retain overall responsibility for the conduct of research; this includes obtaining informed consent from the participants. They must ensure that any person delegated responsibility to participate in the informed consent process is duly authorized, trained and competent to participate according to the ethically approved protocol, Principles of Good Clinical Practice (GCP) and Declaration of Helsinki.

At baseline visit (V0), potential participants will be provided with a verbal explanation of the study and practical details, accompanied by a written patient information sheet stating that the investigation poses minimal risk, involves the collection of a small amount of blood (10 ml) and personal information on a questionnaire. Once eligibility has been established and consent formed signed, procedures will be completed (venipuncture and questionnaires). Subjects will be booked for the next follow-up visit.

### Data protection and patient confidentiality

All investigators and study site staff will comply with the requirements of the Data Protection Act 2018 and GDPR, with regards to the collection, storage, processing and disclosure of personal information. Participant confidentiality will be maintained throughout the investigation. In particular:

- The paper clinical record files/ data will be stored in a locked cabinet
- Each participant will be allocated a unique study identification number. The clinical record will have no identifying patient information on (Patient ID/ Name/ Address/ Date of Birth). The patients will be referred to by their unique study identification number instead
- Participants’ personal details will not be attached to their research results and the decoding list will only be available to a limited number of members of the research team
- All information that is obtained during the study procedures will be treated as private and confidential
- Only the researchers involved in the study will have access to the participants’ data relating to the research project
- Analysis of the data will be undertaken by independent researchers at University of Catania. The study data will all be anonymized and participants will be referred to only by their unique study Identification Number. Published results will only present aggregated, anonymous results and no individuals will be identifiable.

## Results

A total of 1785 subjects have been enrolled in the study. Data cleaning and analyses are ongoing.

## Discussion

This project is the first population-based study that uses seroprevalence data and an objective assessment of the current smoking status in order to examine the association between smoking and SARS-CoV-2 infection susceptibility and severity. Additionally, the study will examine for the first time the magnitude of the seroconversion response in current smokers, compared to former and never smokers as well as the changes in antibody titers over time according to the smoking status. Instead of focusing on hospitalized patients only, the study will also include participants who infected individuals who were either asymptomatic or had COVID-19 but were not hospitalized. It will also consider confounding factors in the association between smoking and SARS-CoV-2 that were not addressed in previous research. Finally, the results from this survey may serve as a reference to other contexts and provide actionable metric to quantify and offer a clear overview of SARS-CoV-2 spread, and the methodology and findings can be exploited in public health research and policies to minimize the disease impact and implement the most appropriate prevention measures to protect susceptible population.

## Data Availability

The data that support the findings of this study are available from the corresponding author, RP, upon reasonable request.

## Acknowledgements

The authors would like to thank Jas Mantero (European Centre for Disease Prevention and Control) and Mark Smolinski (Ending Pandemics) for the constructive feedback about the protocol. The authors would also like to thank Concita Santoro, Annemarie Linder, the Mayor of Troina, Fabio Venezia, Chief of Staff of Protezione Civile, Alessandro Nasca, and all the volunteers of Protezione Civile for their help and assistance.

---

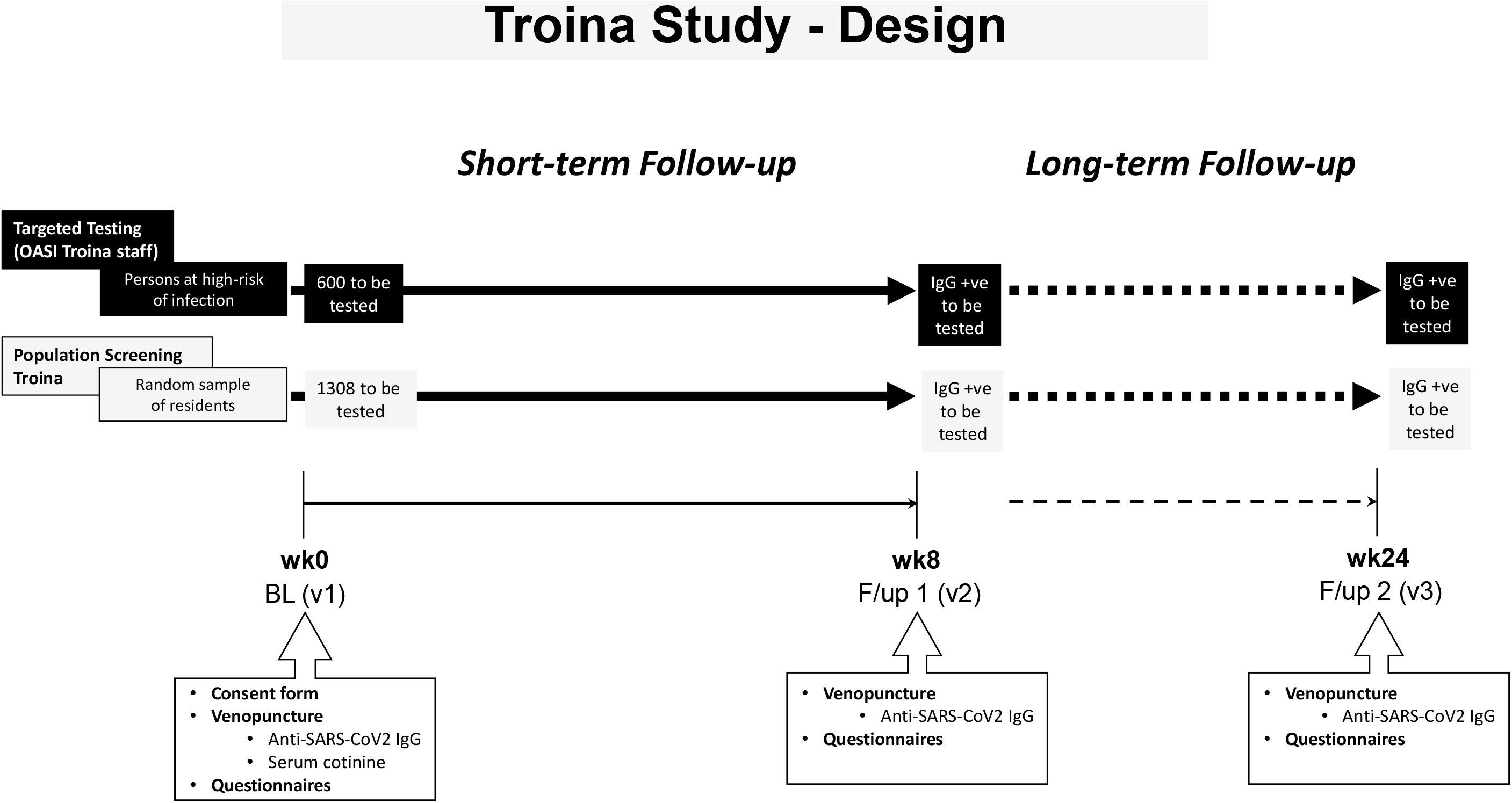

